# Comparative evaluation of case studies and role-playing techniques in enhancing problem-based learning outcomes in Medical Biochemistry education- An educational randomized controlled trial

**DOI:** 10.64898/2026.07.27.26359062

**Authors:** Charushila Y. Kadam, Archana Dhok, Kavneet Khanna, Tripti Waghmare

## Abstract

**Background and objective:** Problem-based learning (PBL) strategies such as case studies and role-playing are increasingly adopted in medical education to promote critical thinking, communication, and teamwork. However, comparative evidence regarding their effectiveness in medical biochemistry remains limited. This study aims to evaluate and compare the impact of case studies and role-playing on learning outcomes, engagement and skill development among first year MBBS students.

**Material and methods:** An educational randomized controlled trial (RCT) with a within-subjects crossover design was conducted in the Department of Biochemistry involving forty-five first-year MBBS students. Participants were randomly assigned to receive case studies and role-playing in a counterbalanced sequence. Learning outcomes were assessed using pre- and post-tests, a structured feedback questionnaire and qualitative reflections. Data were analysed using appropriate parametric and non-parametric tests, along with adjusted analysis using ANCOVA to account for baseline performance.

**Results:** All participants completed both methods, with no significant difference in baseline knowledge between arms (P=0.12). Both case studies and role-playing produced significant pre- to post-test improvements (P<0.001). Unadjusted analyses showed higher learning gains with role-playing, but pre-crossover ANCOVA demonstrated comparable effectiveness after baseline adjustment. Exploratory pooled analysis suggested a moderate cumulative advantage for role-playing. Student feedback indicated higher engagement and perceived skill development with role-playing, while both methods were viewed as relevant and educationally valuable.

**Conclusion:** Both case studies and role-playing were effective PBL strategies for improving understanding and application of medical biochemistry concepts. Although student feedback and unadjusted analyses indicated higher engagement with role-playing, adjusted analysis at initial exposure showed comparable effectiveness between the two methods. Role-playing supported interactive learning and AETCOM-related skills, while case studies strengthened structured analytical reasoning. A combined approach may therefore offer a balanced and comprehensive PBL framework in medical education.

## Introduction

Active teaching methodologies, such as role-play (RP), problem-based learning (PBL), case-based learning (CBL), and team-based learning (TBL), have been increasingly employed to foster critical thinking, communication skills, and teamwork among undergraduate medical students worldwide [1–3]. These methods aim to create a dynamic learning environment, enhancing student engagement, academic satisfaction, and clinical skills [2]. PBL is a well-established teaching approach that integrates basic medical sciences with clinical scenarios, enabling students to actively apply knowledge to real-life situations [3]. PBL has been shown to outperform conventional teaching methods by promoting conceptual learning, critical thinking, and self-directed study while fostering collaborative learning among students [3, 4]. Biomedical science education has also benefited from the incorporation of PBL methodologies. A redesigned curriculum integrating PBL activities, including case studies in haematology and clinical biochemistry, demonstrated improved student engagement, confidence, and performance in case-based assessments. This highlights the potential of PBL to elevate learning experiences and outcomes in applied biomedical sciences [5–7].

Case studies are a classic PBL tool that immerses students in detailed, contextualized scenarios. They require learners to analyse specific situations, apply biochemical concepts, and propose solutions or interventions based on their analysis. This method promotes higher-order thinking by challenging students to integrate knowledge from various domains and apply it to realistic problems. Studies have shown that case studies enhance critical thinking and problem-solving skills, as students engage deeply with the material and practice applying theoretical concepts to practical situations [8]. CBL effectively bridges theory and practice by using real-world cases in healthcare education. Its impact ranges from improved knowledge retention to measurable improvements in patient care [9, 10]. In the realm of dental education, combining case-based learning with role-play was found to significantly improve both critical thinking and teamwork skills. The progressive improvement in student performance across multiple sessions highlights the efficacy of such integrated strategies in clinical learning environments [11].

Role-playing is another PBL strategy where students assume specific roles in simulated scenarios to explore complex concepts and practice interpersonal skills [12]. In role-playing, students act out situations related to biochemical problems, often involving interactions with “patients” or other stakeholders. This method not only helps students understand biochemical principles in a practical context but also develops their communication and teamwork skills. Role-playing can make abstract concepts more tangible and foster empathy and practical problem-solving abilities by immersing students in realistic simulations [13]. A comparative study conducted on second-year undergraduate medical students demonstrated that while debate and role-play were equally effective in improving communication skills, role-play was superior in enhancing knowledge integration and reflecting real-life experiences [1].

While both case studies and role-playing are designed to engage students actively and promote deeper learning, they differ significantly in their approach and execution. Case studies provide a structured, detailed examination of specific scenarios, while role-playing introduces a dynamic, interactive element where students must react in real-time to simulated situations [11, 13, 14]. The effectiveness of these methods can vary based on the learning preferences of students, the complexity of the content, and the specific educational goals. Previous research has explored the individual impacts of these methods on learning outcomes. However, there is a paucity of studies directly comparing the efficacy of case studies and role-playing within the context of Biochemistry education.

Understanding how each method influences students’ comprehension of Biochemistry concepts, engagement levels, and overall learning outcomes is crucial for optimizing teaching strategies. The present study was thus undertaken to evaluate and compare the efficacy of case studies and role-playing in enhancing PBL outcomes in Biochemistry education and determine the more effective PBL method. Addressing this gap would offer evidence-based recommendations for educators, enabling them to design more effective and engaging learning environments that enhance student learning experiences and outcomes in medical Biochemistry education. These findings carry practical implications for curriculum development, equipping educators to create learning strategies that better prepare students for their future medical careers.

## Material and methods

The study was conducted in the Department of Biochemistry, Sukh Sagar Medical College and Hospital, Jabalpur, over a duration of six months. The study was approved by Institutional Ethics Committee (IEC) of Sukh Sagar Medical College and Hospital [Ref No. SSMCH/Ethics/App/2024/5143]. All research activities were conducted in compliance with the ethical guidelines and standards outlined by the IEC. Participants were prospectively recruited for this study between 13 December 2024 and 22 December 2024. Prior to participation, all students were provided with detailed information about the study, including its purpose, procedures, potential risks, and benefits. They were informed that their participation was voluntary, and they could withdraw from the study at any time without any consequences. Informed written consent forms were obtained from all participants, ensuring they understand the nature of the study and agree to participate. The consent was documented and securely stored in digital format. This study did not include any minors, and therefore, parental or guardian consent was not applicable. This randomized controlled trial (RCT) was conducted as an educational intervention study. At the time of study initiation, prospective trial registration was not mandated for non-clinical interventions.

An educational Randomized Controlled Trial (RCT) with a within-subjects crossover design was utilized, wherein all participants were experienced both case studies and role-playing interventions in a counterbalanced order to mitigate order effects. The study participants were first-year MBBS students, with inclusion criteria comprising Phase I MBBS students attending Biochemistry sessions and exclusion criteria excluding those who were previously exposed to the proposed interventions. Participants were randomly assigned to either Group A (case studies first) or Group B (role-playing first). A priori sample size estimation for the within-subject crossover design was performed using assumptions for a paired comparison. Assuming a medium effect size (d=0.5), a two-tailed significance level of 5% (α=0.05), and a statistical power of 80% (β=0.20), the minimum required sample size was estimated to be 32 participants. To account for an anticipated attrition rate of 20%, the target sample size was increased to 39 participants. However, to ensure equitable educational opportunities and avoid excluding students from either instructional intervention, all eligible first year MBBS students (n=150) were invited to participate in the study.

### Enrolment, randomization and allocation concealment

A total of 150 first-year MBBS students were initially enrolled for participation. Simple randomization was performed using a lottery method. An independent faculty member, who had no role in recruitment, consent procedures, or data collection, prepared 150 identical, opaque, folded slips, marked in equal numbers (75 each) with either ‘A’ (case studies → role playing) or ‘B’ (role playing → case studies). The slips were thoroughly mixed and placed in a covered container. After providing informed consent, each student randomly drew one slip, and the group assignment was recorded. This process ensured adequate randomization and allocation concealment preventing both investigators and participants from knowing the assignment sequence in advance.

Of the 150 students who underwent randomization, 105 students were classified as drop-outs because they did not attend the scheduled PBL intervention sessions and therefore did not meet the minimum participation requirement. Consequently, 45 students who completed all intervention sessions and assessments were retained in the study and included in the final complete-case analysis according to their originally assigned groups (Group A:22 and Group B: 23).

A CONSORT-style flow diagram (Fig. 1) illustrates the process of enrolment, randomization, allocation, attendance and analysis, providing a clear, visual summary of participant progression through-out the study.

**Fig. 1:**
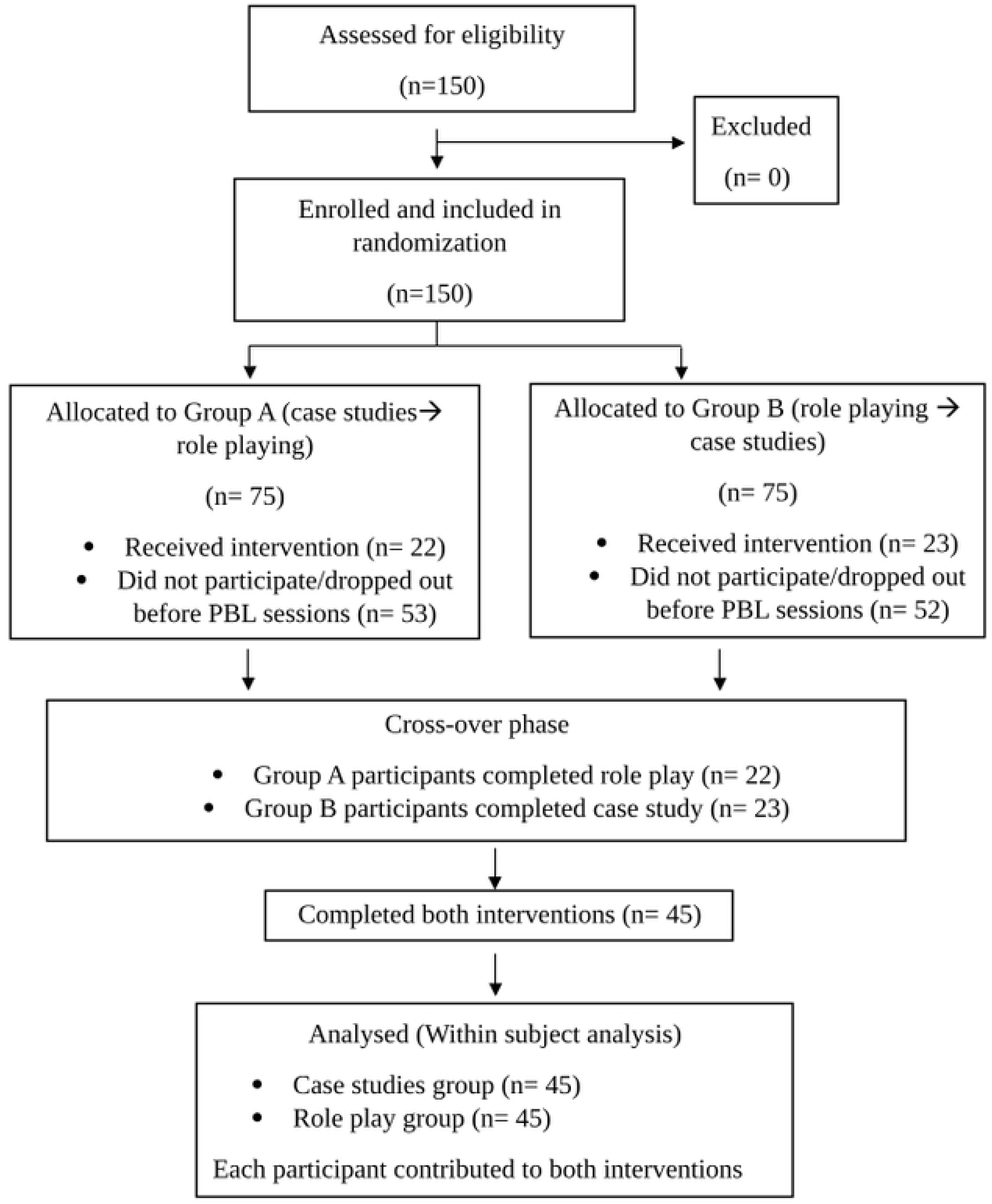
CONSORT-style flow diagram for within-subject randomized crossover trial depicting enrolment, randomization to initial instructional arm, crossover to the alternate method, session-wise participation and inclusion in the final analysis.

### Data Collection Tool

To accurately assess the impact of case studies and role-playing on students’ understanding and engagement in Biochemistry, multiple data collection tools were employed:

#### 1. Pre- and Post-Tests

Designing: Pre- and post-tests were developed to evaluate students’ knowledge of Biochemistry concepts before and after each intervention (case studies and role-playing). These tests were consisted of multiple-choice questions (MCQs), short-answer questions, and scenario-based questions that reflect the key learning objectives of the course.

Validation: To ensure content validity, the test items were reviewed by a panel of Biochemistry educators and curriculum experts. The items were evaluated for clarity, relevance, alignment with learning objectives and cognitive level. Based on the feedback, modifications were incorporated to refine item wording, difficulty level and content alignment.

Reliability assessment: Within-study reliability of the test instruments was evaluated by calculating Kuder-Richardson Formula-20 (KR-20) and Cronbach’s α for pre-test and post-test scores. Internal consistency was examined using dichotomous scoring for objective knowledge items ensuring that the instrument consistently measured the intended constructs.

#### 2. Student Engagement and Satisfaction Questionnaire

Designing: A Likert scale-based questionnaire was designed to measure students’ engagement and satisfaction with each PBL method. The questionnaire included items related to perceived understanding, interest level, interaction quality, and overall satisfaction with the learning experience.

Validation: The questionnaire was subjected to expert review for face and content validity.

Reliability assessment: Internal consistency reliability of the polychotomous (Likert scale) feedback questionnaire was assessed during the study using Cronbach’s α, to ensure reliability of the responses across questionnaire items.

#### 3. Qualitative Feedback

Designing: Open-ended questions were included in the feedback questionnaire to gather anonymous qualitative feedback on students’ experiences with case studies and role-playing. To avoid influencing students’ responses during the intervention period, the feedback questionnaire was administered only after completion of all case study and role-playing sessions and their corresponding assessments. This provided additional insights into students’ perceptions of the effectiveness of each instructional method and the challenges encountered during the learning process.

Validation: The open-ended questions were reviewed by educational experts to ensure clarity, neutrality, and their ability to elicit meaningful responses. The responses were analysed descriptively using keyword-based percentage analysis to summarize recurring perceptions and experiences rather than formal thematic analysis.

### Data collection procedure

#### Interventions

The study employed two problem-based learning approaches: case studies and role-playing. The case-study sessions included four clinical scenarios based on acute pancreatitis, rickets, beriberi, and pellagra whereas the role-playing sessions included four scenarios based on myocardial infarction, night blindness, scurvy, and macrocytic anemia. All topics were selected from the National Medical Commission (NMC) Competency-Based Medical Education (CBME) curriculum and focused on the application of biochemical principles in clinically relevant situations. To minimize content-related bias, the scenarios were designed to be comparable in curricular importance, educational objectives, cognitive level, prerequisite knowledge requirements and problem-solving skills. All scenarios required students to apply biochemical concepts to interpret clinical findings, understand disease mechanisms and support diagnosis. The comparability of the scenarios was reviewed by faculty members from the Department of Biochemistry.

#### Procedure

Before each instructional session, participants completed a pre-test to assess their baseline knowledge of the relevant topic. Students assigned to Group A initially participated in the case-study sessions followed by the role-playing sessions, whereas those assigned to Group B received the interventions in the reverse order. Following each session, participants completed a post-test to evaluate knowledge acquisition and conceptual understanding. A counterbalanced crossover design was employed to minimize order effects, ensuring that all participants experienced both instructional methods in different sequences.

#### Baseline characteristics and equal exposure

As this study employed a within-subject randomized controlled crossover design, all participants (n=45) were exposed to both instructional methods. Demographic variables such as age and gender were not used for group allocation because all students eventually received both interventions, ensuring equivalent demographic exposure. Baseline comparability between the two randomized sequences was assessed using pooled pre-test scores from all four sessions to obtain a robust estimate of baseline knowledge.

#### Exposure across arms

Each session of case study and role play was conducted for an identical duration of 60 minutes ensuring uniform time-on-task across both the methods. All sessions were facilitated by the same faculty members. To minimize facilitator related variability, facilitators adhered to a standardized session handouts and identical instructional guidelines. Assessment procedures were also kept identical across arms. Together, these measures ensured that any observed differences in learning outcomes were attributable to the instructional methods themselves rather than differences in exposure, delivery or assessment conditions.

### Statistical analysis

Prior to inferential analysis, the normality of pre-test scores, post-test scores and score improvement values were assessed using the Shapiro-Wilk test. Depending on data distribution either parametric or non-parametric statistical tests were applied. Within group comparisons of pre-test and post-test scores were performed using the paired Student’s t-test for normally distributed data and the Wilcoxon signed-rank test for non-normally distributed data. Baseline comparability between the case study and role play arms was assessed by comparing pooled pre-test scores using the Mann-Whitney U test. Comparisons of post-test scores and score improvements between the study groups were conducted using the unpaired t-test or Mann-Whitney U test, as appropriate. As the primary adjusted analysis, analysis of covariance was performed to determine whether learning outcomes differed between the study groups after adjusting for baseline academic performance, with post-test score as the dependent variable, group allocation as the independent variable and pre-test score as the covariate. The association between score improvements achieved during case study and role-playing sessions was evaluated using Spearman’s rank correlation coefficient. Student feedback was analysed using descriptive statistics and percentage analysis. Responses to Likert-scale items were expressed as percentages for each response category (strongly agree, agree, neutral, disagree and strongly disagree) to facilitate comparison between the two instructional methods. Open ended feedback was reviewed descriptively and percentage analysis of recurring key words was performed to assess student engagement and satisfaction. A p-value of <0.05 was considered statistically significant.

## Results

A total of 150 first-year MBBS students were enrolled and randomized; 45 participants completed all study procedures and were included in the final complete-case analysis. Table 1 depicts the demographic and baseline characteristics of the participants. As all students eventually received both instructional methods-case studies and role-playing-demographic variables were inherently equivalent across arms. Baseline knowledge comparability was assessed using pooled pre-test scores from all four sessions of case studies and role-play arms, reflecting overall equivalence prior to instruction. The Mann-Whitney U test indicated no statistically significant difference in baseline pre-test scores between two instructional arms (Z=1.52, P=0.12). The associated effect size was very small (r=0.08), confirming that both groups were comparable with minimal practical difference between the groups prior to the intervention. As session 1 represented the pre-crossover phase, adjusted between arm comparison using ANCOVA was restricted to session 1 to preserve the randomized structure of the trial.

**Table 1:** Demographic and baseline characteristics of the participants.

| Characteristics | Value |
| --- | --- |
| **Gender, n (%)** |  |
| Male | 7 (15.55%) |
| Female | 38 (84.44%) |
| **Age (years)** |  |
| Mean (SD) | 19.95 (1.38) |
| Range | 18-24 years |
| **Baseline knowledge comparison** |  |
| Pre-test scores (case studies), Median (IQR) | 7 (5) |
| Pre-test scores (case studies), Mean (SD) | 7.13 (3.92) |
| Pre-test scores (role-play), Median (IQR) | 6 (5) |
| Pre-test scores (role-play), Mean (SD) | 6.52 (3.78) |
| Mann-Whitney U | 17696.5 |
| p-value | 0.12 (Not significant) |
| Z | 1.52 |
| Effect size (r) | 0.08 |

### Reliability analysis

The internal consistency reliability of the knowledge assessment instruments was evaluated using KR-20/Cronbach’s α. For dichotomously scored items. Because individual session-wise tests contained a limited number of items, reliability was estimated using pooled pre-test and post-test data across the four case studies sessions and four role playing sessions to obtain more stable estimates. The pooled pre-test instruments demonstrated acceptable internal consistency (case study: Cronbach’s α /KR20= 0.66; role playing: Cronbach’s α /KR20= 0.67), while the pooled post-test case study instrument demonstrated good reliability (Cronbach’s α /KR20=0.78). The pooled post-test role playing instrument showed lower internal consistency (Cronbach’s α /KR20=0.52). The relatively lower reliability observed in the role-play post-test may reflect reduced variance in responses following instruction, consistent with a ceiling tendency commonly observed in post-intervention assessments rather than poor item construction. The relatively low mean inter-item correlations observed for the pre-test and post test instruments in both the case study and role-playing sessions reflect the multi-domain, heterogenous nature of the knowledge assessment, a feature commonly seen in educational tests and not indicative of compromised overall reliability (Supplementary Table S1). The student feedback questionnaire demonstrated excellent internal consistency (Cronbach’s α=0.87). Detailed reliability statistics are provided in Supplementary Table S1.

### Normality assessment

The normality of pre-test scores, post-test scores and score improvement values was assessed using the Shapiro-Wilk test. The data demonstrated a mixed distribution, with some variables satisfying normality assumptions and others showing significant departures from normality. Accordingly, parametric tests were applied to normally distributed variables, whereas non-parametric tests were used when normality assumptions were violated. Detailed results of the normality assessment are presented in Supplementary Table S2.

The pre-test and post-test performance for the case study and role play intervention is summarized in table 2 and table 3 respectively. As the score distributions violated normality assumptions for several sessions, results are primarily presented as median and interquartile range (IQR), with mean (SD) provided as supplementary descriptive statistics. Fig. 2A and Fig. 2B illustrates the session-wise pre-test and post-test score distributions for the case study and role-play arms, respectively. The analysis demonstrated a statistically significant improvement in post-test scores compared with pre-test scores for both instructional methods, indicating that both case studies and role-playing were effective in enhancing students’ learning outcomes.

**Fig. 2A:**
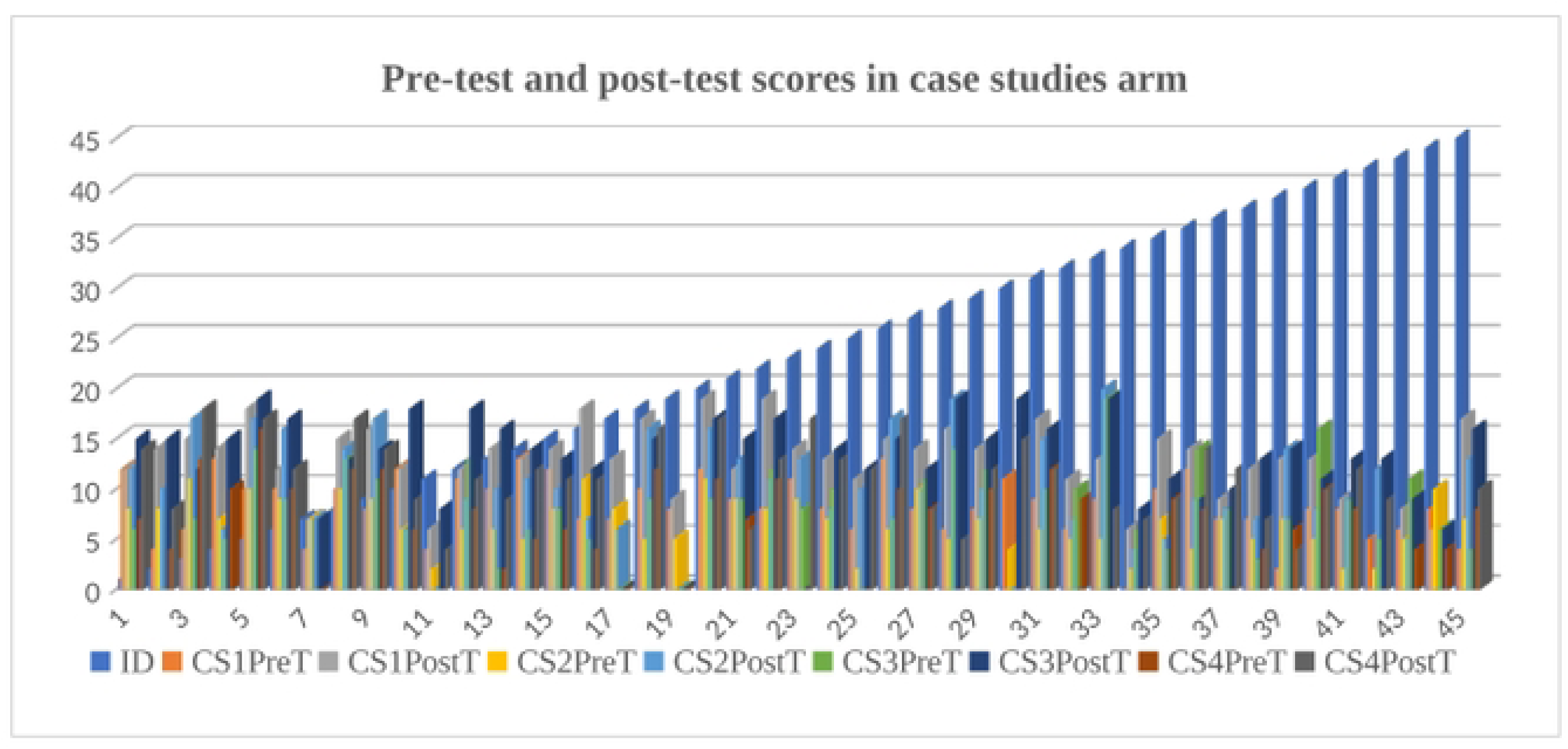
Session-wise distribution of pre-test and post-test scores for the case study arm Pre-test and post-test scores in case studies arm.

**Fig. 2B:**
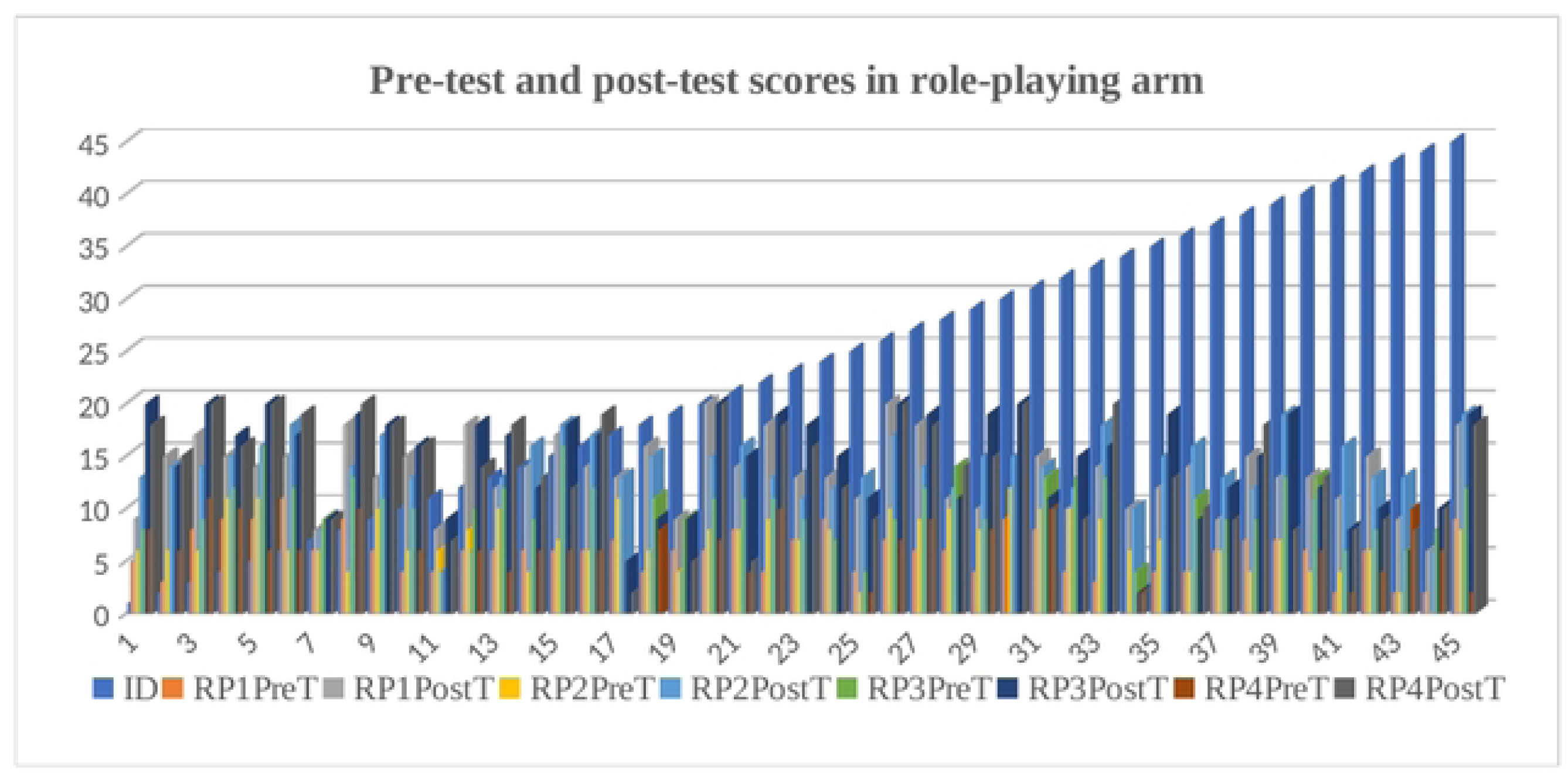
distribution of pre-test and post-test scores for the role-play arm. Pre-test and post-test scores in role-playing arm.

**Table 2:**
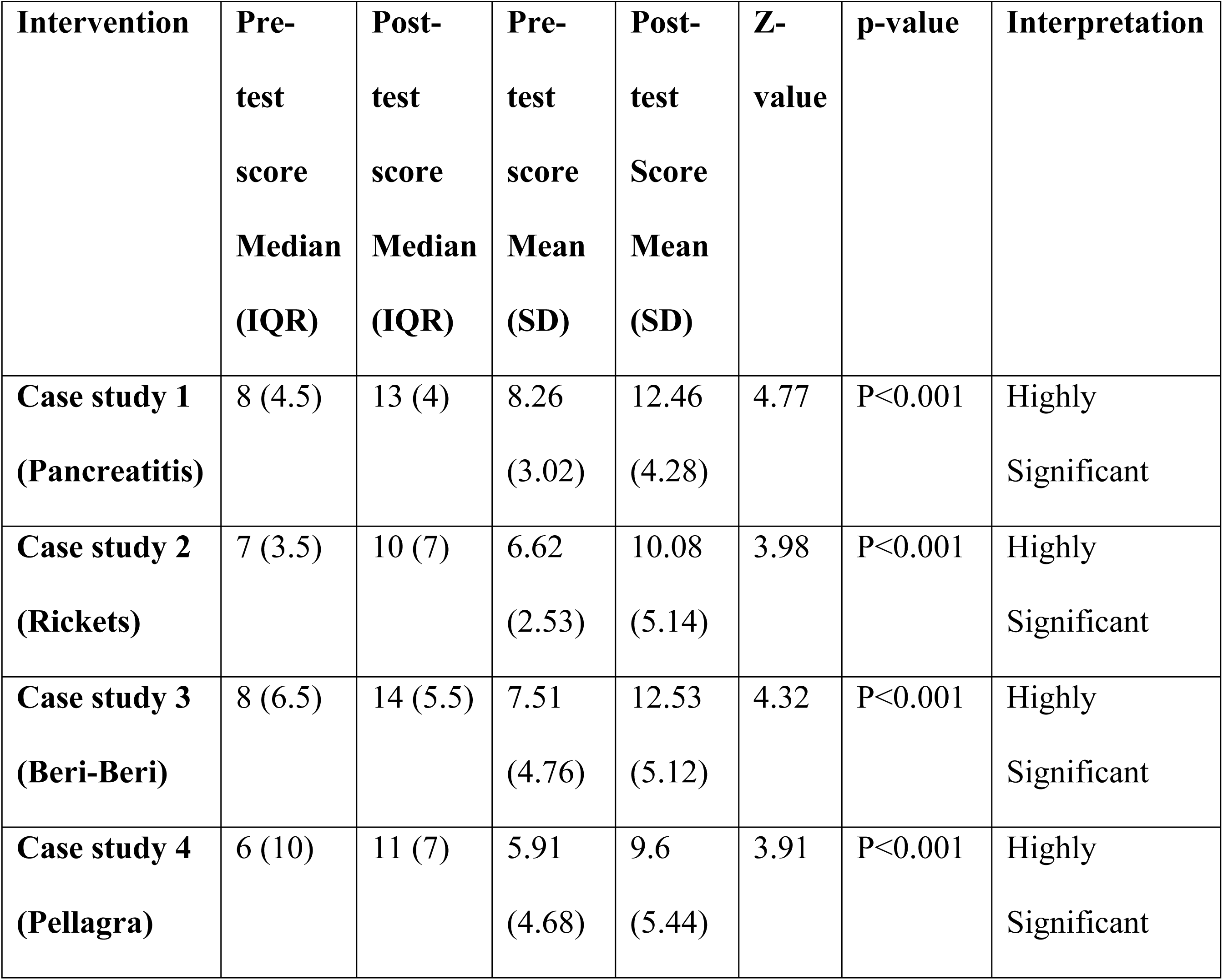
Within group comparison of pre-test and post-test scores for case study sessions.

**Table 3:** Within group comparison of pre-test and post-test scores for role-play sessions.

| **Intervention** | **Pre-test score**<br>**Median**<br>**(IQR)** | **Post-test score**<br>**Median**<br>**(IQR)** | **Pre-test score**<br>**Mean**<br>**(SD)** | **Post-test score**<br>**Mean**<br>**(SD)** | **Z-value**<br>**/t-value** | **p-value** | **Interpretation** |
| --- | --- | --- | --- | --- | --- | --- | --- |
| **Role-play 1**<br>**(Myocardial infarction)** | 6 (3) | 14 (4.5) | 5.8<br>(2.30) | 13.15<br>(3.90) | $t=12.4$ | $P<0.001$ | Highly Significant |
| **Role-play 2**<br>**(Night blindness)** | 7 (3) | 14(4) | 6.93<br>(2.69) | 13.51<br>(3.57) | $Z=5.70$ | $P<0.001$ | Highly Significant |
| **Role-play 3**<br>**(Scurvy)** | 10 (4) | 16 (8.5) | 9.2<br>(4.20) | 14.53<br>(4.79) | $Z=5.17$ | $P<0.001$ | Highly Significant |
| **Role-play 4**<br>**(Macrocytic anemia)** | 4 (7) | 14 (9) | 4.17<br>(3.78) | 13.17<br>(5.60) | $Z=5.48$ | $P<0.001$ | Highly Significant |

Effect size analysis for within group pre-test to post-test comparisons in the case study sessions demonstrated large effects across all four sessions (r=0.74 for case study 1, 0.62 for case study 2, 0.68 for case study 3 and 0.64 for case study 4), indicating substantial learning gains following the case study intervention.

Effect sizes for within group pre-test to post-test comparisons in the role-playing sessions indicated large to very large learning effects. For role-play session 1, the magnitude of improvement was quantified using Cohen’s d and demonstrated a very large effect (d=1.85). For role-play sessions two, three and four, effect sizes were calculated using the non-parametric effect size (r), yielding consistently large effects (r=0.86, r=0.80 and r=0.84, respectively). Collectively, these findings indicate substantial learning gains following the role-playing intervention across all sessions.

Between arm comparisons of post-test scores (Table 4) and score improvements (Table 5) were performed using the appropriate statistical tests based on data distribution. The pooled analyses were performed by combining scores from all four sessions within each instructional method to estimate the overall difference in post-test performance and learning gains (Table 6). These analyses complemented session-wise comparisons and provided a global assessment of instructional effectiveness across the study period. As these comparisons do not adjust for baseline scores, and include post-crossover data, they were interpreted as descriptive and comparative rather than causal.

**Table 4:** Between arm comparison of post-test scores for case studies and role-playing sessions.

| Session | U | Z-value | p-value | Interpretation |
| --- | --- | --- | --- | --- |
| Case study 1 vs Role play 1 | 932.5 | -0.64 | $p=0.51$ | Not significant |
| Case study 2 vs Role play 2 | 597.5 | -3.35 | $P<0.001$ | Highly Significant |
| Case study 3 vs Role play 3 | 766.5 | -1.98 | $p=0.04$ | Significant |
| Case study 4 vs Role play 4 | 646 | -2.96 | $p=0.003$ | Highly Significant |

**Table 5:**
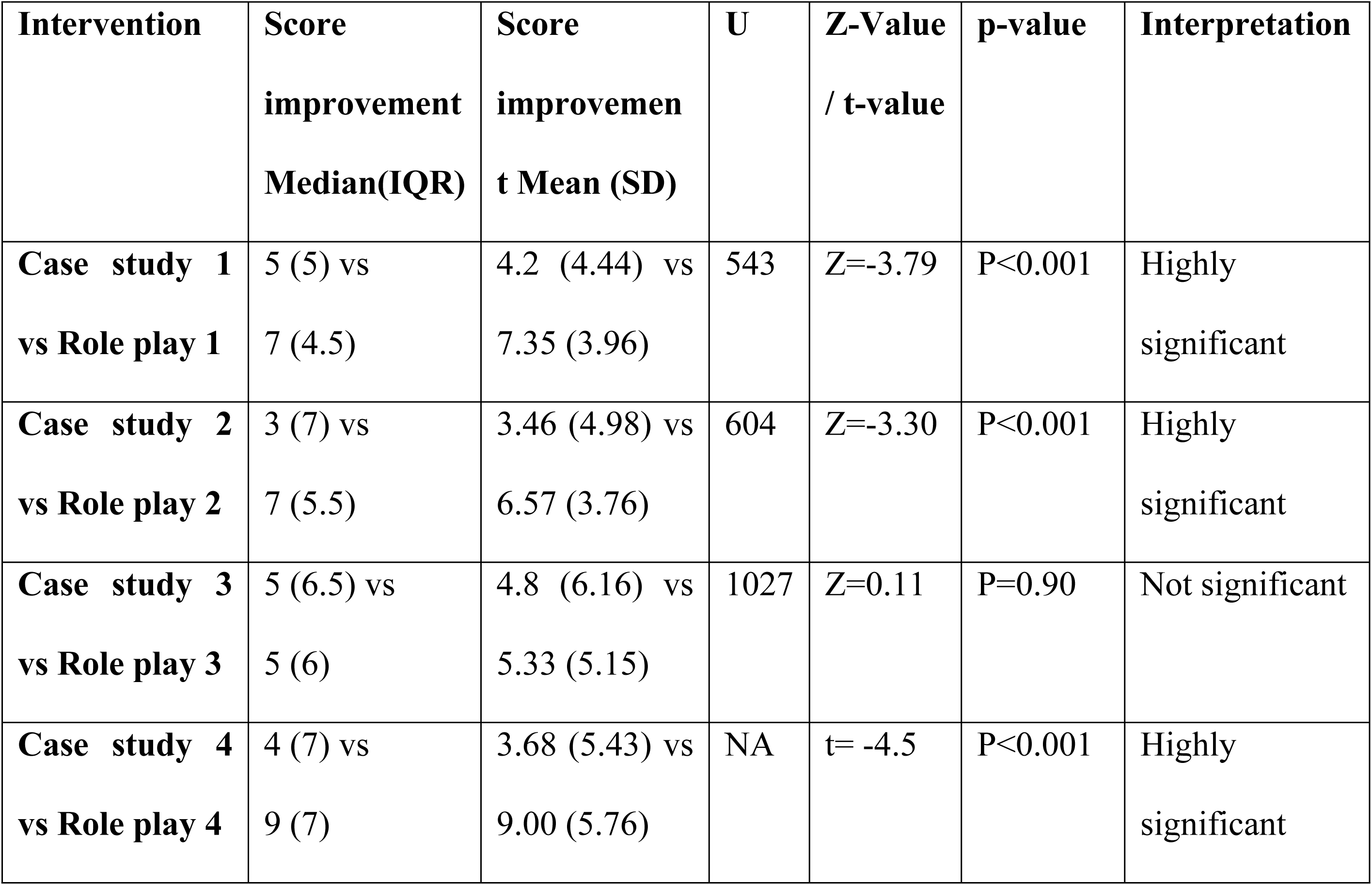
Between arm comparison of score improvement in scores in case studies and role-playing sessions.

| Intervention | Score improvement<br>Median(IQR) | Score improvement<br>t Mean (SD) | U | Z-Value<br>/ t-value | p-value | Interpretation |
| --- | --- | --- | --- | --- | --- | --- |
| Case study 1<br>vs Role play 1 | 5 (5) vs<br>7 (4.5) | 4.2 (4.44) vs<br>7.35 (3.96) | 543 | Z=-3.79 | P<0.001 | Highly<br>significant |
| Case study 2<br>vs Role play 2 | 3 (7) vs<br>7 (5.5) | 3.46 (4.98) vs<br>6.57 (3.76) | 604 | Z=-3.30 | P<0.001 | Highly<br>significant |
| Case study 3<br>vs Role play 3 | 5 (6.5) vs<br>5 (6) | 4.8 (6.16) vs<br>5.33 (5.15) | 1027 | Z=0.11 | P=0.90 | Not significant |
| Case study 4<br>vs Role play 4 | 4 (7) vs<br>9 (7) | 3.68 (5.43) vs<br>9.00 (5.76) | NA | t= -4.5 | P<0.001 | Highly<br>significant |

**Table 6:**
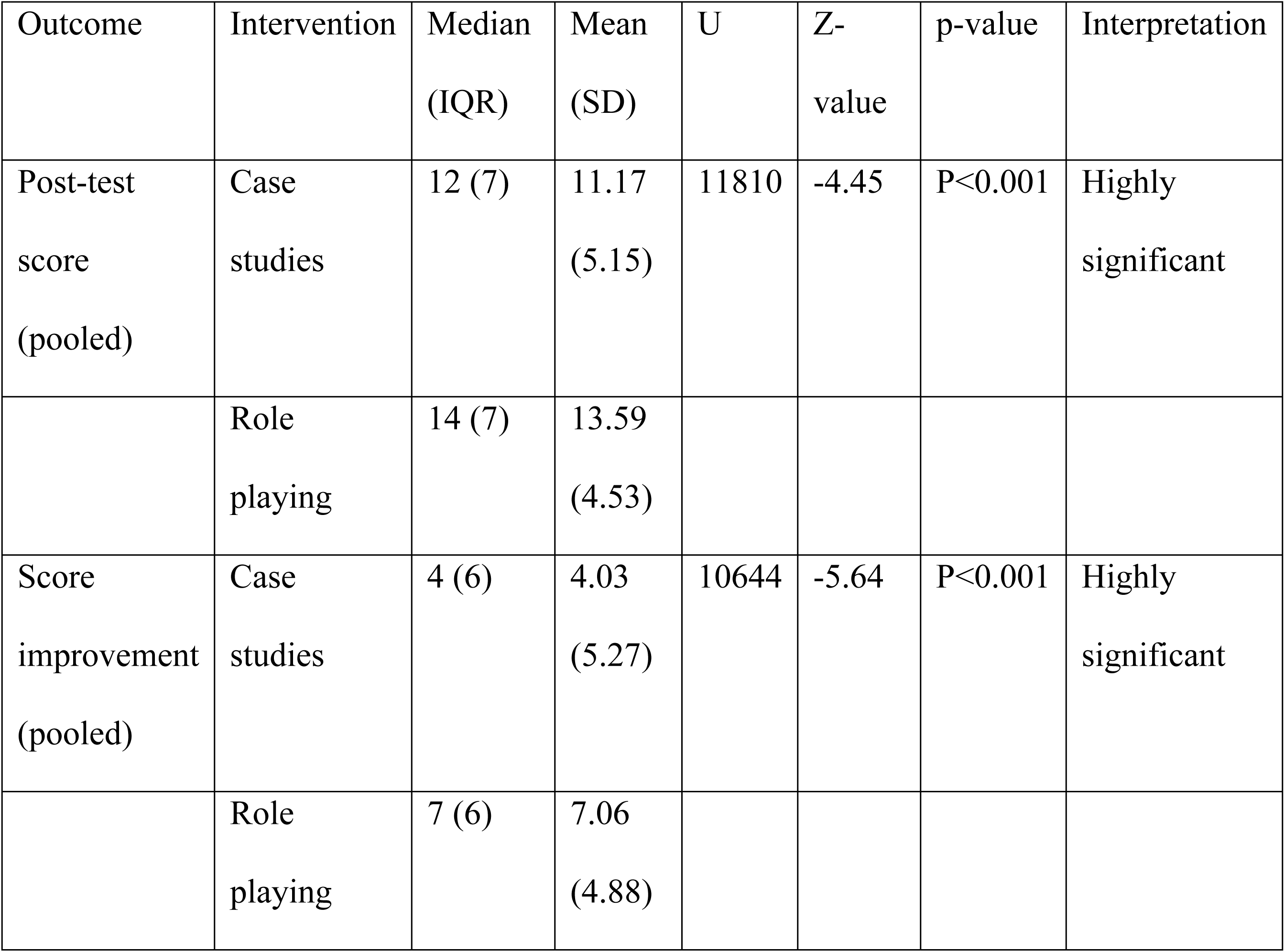
Pooled between-arm comparison of post-test scores and score improvement.

Descriptive statistics for post-test scores are presented in table 2 and table 3 whereas between arm comparisons are summarized in Table 4. Effect size analysis for between arm comparisons for post-test scores demonstrated that the effect size for session 1 was negligible (r=0.068), consistent with the non-significant difference observed between the two instructional methods. In contrast, session 2 demonstrated a moderate effect size (r=0.35), indicating a meaningful advantage of role-playing instructional method over the case studies. Session 3 and 4 showed small to moderate effect sizes (r=0.21 and r=0.31, respectively), suggesting modest but statistically significant differences in post-test performance between the case study and role-playing arms. Overall, these findings indicated that while some sessions exhibited minimal differences, others demonstrated educationally relevant advantages between instructional strategies.

Effect size analysis of between arm differences in score improvements showed a moderate effect favouring role-playing in session 1 (r=0.40) and session 2 (r=0.35), indicating meaningful educational advantages of role playing over case studies in these sessions. In contrast, session 3 demonstrated a negligible effect size (r=0.012), consistent with the absence of a statistically significant difference in score improvement between the two instructional methods. For session 4, a large effect size was observed (Cohen’s d=0.95), indicating a substantial improvement in learning gains with role-playing compared to case studies. Furthermore, results of Spearman’s rank correlation analysis of score improvements between case studies and role-playing sessions revealed a significant medium positive correlation (r=0.30, P<0.001). This finding indicated that participants who showed greater learning gains in one instructional method also tended to achieve greater gains in the other, suggesting consistency in individual learning responsiveness across instructional methods rather than superiority of a single approach.

The effect size analysis for pooled comparison of post-test scores between case studies and role-playing sessions demonstrated a small-to-moderate effect size (r=0.23), indicating that role-playing was associated with modestly higher overall post-test performance compared with case studies. Similarly, the pooled comparison of score improvement showed a moderate effect size (r=0.30), suggesting that role-playing produced greater overall learning gains than case studies when data from all four sessions were considered together.

### Adjusted between-arm comparison using ANCOVA

#### Primary adjusted analysis (Session 1)

To address baseline differences and leverage the randomized design, a one-way analysis of covariance (ANCOVA) was conducted for session 1 post-test scores, with instructional arm as the independent variable and corresponding pre-test scores as the covariate. Session 1 was selected as the primary analysis because it occurred prior to crossover, ensuring that participants were naïve to the alternate instructional method. After adjusting for pre-test scores, there was no statistically significant difference in post-test performance between the case study and role-playing arms, F (1, 42) =1.28, p=0.26, partial η² = 0.030, indicating a small effect size. The covariate pre-test score, was not significantly associated with post-test performance, F (1, 42) = 1.96, p= 0.16. These findings indicate that both instructional methods were comparably effective at initial exposure.

#### Exploratory adjusted analysis

An exploratory ANCOVA was subsequently performed on pooled post-test scores across all sessions to examine overall instructional effects while adjusting for pooled pre-test scores. After adjustment, a statistically significant difference was observed between instructional arms, F (1, 357) = 30.35, p<0.001, partial η² = 0.078, indicating a moderate effect size, with instructional arm accounting for approximately 7.8% of the variance in post-test scores. Adjusted mean post-test scores were higher for the role-playing arm compared with the case study arm. Given the crossover design and repeated observations per student, this pooled ANCOVA was considered exploratory and was interpreted as reflecting cumulative learning trends rather than a definitive causal effect.

In addition to the quantitative outcome measures, percentage analysis was performed to assess and compare the feedback for each problem-based learning (PBL) method employed in the study. The general feedback on the case studies revealed that a majority of students (91.1%) agreed or strongly agreed that the case studies were relevant to the Biochemistry curriculum. Only 8.8% were neutral, with no one disagreeing. This suggests high relevance to the course content. Moreover, 73.32% of students either strongly agreed or agreed that case studies helped deepen their understanding of Biochemistry concepts. 26.66% were neutral, indicating that a significant portion of students might not have fully benefitted from case studies in enhancing their understanding. 66.66% found the case studies engaging and thought-provoking, with 31.11% neutral. A small minority (2.22%) disagreed, which suggests that while most students found case studies engaging, there’s a segment that didn’t resonate as much. 71.10% (strongly agree + agree) felt that case studies improved their critical thinking skills, which shows that case studies were effective in fostering analytical thinking. 26.66% were neutral, with 2.22% disagreeing. 64.44% felt that case studies allowed them to effectively apply theoretical knowledge to practical scenarios whereas 28.88% were neutral, and 6.66% disagreed, suggesting that case studies helped most but not all students apply their theoretical knowledge. The general feedback on the role playing revealed that 91.11% of respondents agreed or strongly agreed that role-playing scenarios were relevant to the Biochemistry curriculum. 8.88% were neutral, but no one disagreed, which shows a high degree of relevance for this method as well. Moreover, 91.11% agreed or strongly agreed that role-playing enhanced their ability to think and respond in real-time, indicating this method was effective in improving spontaneous thinking and decision-making skills. 91.10% of students found role-playing engaging and interactive, with only 8.88% neutral, which highlights the high level of student involvement in role-playing. In addition to this, 93.32% of students agreed or strongly agreed that role-playing helped improve communication and teamwork skills. This method has had a significant impact on fostering collaborative and communication skills. 77.76% of students found that role-playing made Biochemistry concepts more tangible and easier to understand whereas 17.77% were neutral, and 2.22% disagreed, showing that while most students found this method helpful, there were a few who did not find it as beneficial. The comparative feedback on the case studies versus role-playing revealed that 51.11% felt role-playing helped them better understand Biochemistry concepts. 40% believed both methods helped equally, while only 8.88% preferred case studies. This suggests role-playing was more effective for understanding the concepts compared to case studies. In addition to that substantial 84.44% students found role-playing more engaging. Only 4.44% preferred case studies, with 11.11% stating both methods were equally engaging. This clearly points to role-playing as the more engaging method. Comparative feedback on improvement in critical thinking and problem-solving skills revealed 60% participants felt that role-playing contributed more to improving their critical thinking and problem-solving skills. 24.44% thought both methods contributed equally, while 15.55% favoured case studies. This further underscore role-playing’s positive impact on problem-solving and critical thinking. Moreover, role-playing was favoured (64.43%) by participants in improving communication skills, especially in the context of attitudes, ethics, and communication (AETCOM) module, a longitudinal component of the NMC CBME curriculum designed to develop professional attitudes, ethical reasoning and communication skills in medical students. Only a small percentage felt case studies contributed more (4.44% combined). A substantial 20% felt both methods contributed equally, while 11.11% didn’t respond. These observations indicate that role-playing can facilitate communication and interpersonal skills, which are critical in AETCOM. In conclusion, student feedback indicated that both case studies and role-playing were perceived as valuable and complementary instructional strategies within the PBL framework. Case studies were consistently regarded as highly relevant to the biochemistry curriculum and were perceived to support conceptual understanding analytical reasoning and application of theoretical knowledge. Role-playing on the other hand, was more frequently associated with higher levels of student engagement, active participation, communication, teamwork and real-time problem solving. While feedback trends suggested a greater perceived impact of role-playing on engagement and communication-related outcomes, these observations reflect student perceptions rather than definitive evidence of instructional superiority. Overall, the feedback reports the integration of both case studies and role-playing within medical biochemistry teaching with case reinforcing structured conceptual learning and role-playing facilitating experiential, interactive skill development. Leveraging both methods in a balanced manner may help optimize learning experiences addressing diverse learning preferences and educational objectives, rather than relying on a single instructional approach.

Additionally, descriptive analysis of the anonymous qualitative feedback using keyword-based percentage analysis indicated that students perceived both instructional methods positively. For the case study method 64.44% of students reported that its engaging nature and the effective real-world scenarios enhanced their understanding of complex concepts and theoretical knowledge, whereas 24.44% identified lengthy sessions and time constraints as the main challenges. For the role-playing method, 37.77% of students reported that it helped them overcome stage fear and improve communication skills, although 15.55% considered nervousness and stage fear to be barriers to participation. A small percentage (4.44%) suggested shortening case study sessions to enhance time management.

Descriptive analysis of the anonymous open-ended reflections further indicated that 48.88% of students perceived both instructional methods as beneficial for future learning and clinical practice by improving communication skills and confidence during patient interactions. In addition, 26.66% of students expressed that the sessions were interactive, fun, and enjoyable, reinforcing the value of these teaching methods in making learning engaging. These descriptive findings complemented the quantitative feedback by providing additional insights into students’ perceptions and learning experiences.

## Discussion

The findings of this study provide valuable insights into the comparative effectiveness of case studies and role-playing as problem-based learning (PBL) strategies in enhancing medical biochemistry education. Overall, the results demonstrate that both instructional approaches were effective in improving student performance, supporting their value as active learning methodologies. These results align with previous studies and offer critical implications for the curriculum design and instructional planning in medical education.

Our analysis revealed a significantly higher post-test score compared to pre-test scores in case studies (P<0.001) as well as role-playing sessions (P<0.001) indicating that both structured PBL activities were effective in improving student performance, understanding and application of biochemistry concepts. These findings are consistent with prior educational research demonstrating that interactive learning strategies promote deeper cognitive engagement and improved academic outcomes [15, 16]. Moreover, unadjusted session-wise and pooled analysis revealed higher post-test scores and greater score improvements during several role-playing sessions compared with the case studies. Similarly, pooled rank-based effect size estimates suggested small-to-moderate cumulative advantages favouring role-playing when data from all sessions were considered together. These observations suggest that role-playing may offer incremental learning benefits with repeated exposure, potentially due to its active, immersive, participatory and experiential nature, which aligns with educational theories favouring experiential learning for deeper comprehension. However, when the randomized structure of the trial was preserved by restricting adjusted analysis to the pre-crossover phase (session 1), the primary ANCOVA did not demonstrate a statistically significant between-arm difference in post-test scores after controlling for baseline performance. This finding indicates that at initial exposure, both instructional methods were comparably effective. In contrast, exploratory pooled ANCOVA analysis suggested a moderate cumulative advantage for role-playing after adjustment for baseline scores. Given the cross-over design and repeated observations per participant, this finding should be interpreted cautiously and viewed as indicative of cumulative learning trends rather than definitive causal superiority. Future studies employing longitudinal analytical approaches, such as mixed effects modelling, may further elucidate time-dependent and cumulative instructional effects while appropriately accounting for within student correlations.

Previous studies provide a useful context for interpreting these findings. Chan et al. [15] demonstrated that incorporating role-plays in PBL improves students’ learning motivation, creativity, and understanding of the patient and family needs emphasizing the value of active and experiential learning. However, their work was focused on enriching traditional PBL rather than directly comparing instructional modalities. To the best of our knowledge, the present study is the first to directly compare the effectiveness of role-playing and case studies in PBL sessions using a randomized controlled trial with a within-subject study design in the context of medical education. Similarly, Sanghani et al. [16], who evaluated the effectiveness of case-based learning (CBL) in teaching clinical biochemistry to Phase-I MBBS students, reported significantly higher post-test scores with CBL compared to interactive didactic lectures (IDL). Their findings align with our results showing significant improvements in post-test performance following both case studies and role-playing sessions. Our study extends their findings by directly contrasting two interactive instructional strategies. Additionally, while Sanghani et al. [16] observed that CBL promoted active participation, teacher-student communication, and knowledge application, our study builds on this by demonstrating that role-playing enhances not only these aspects but also creativity, student autonomy, and the understanding of complex problem-solving in a more immersive, experiential context. Both studies reinforce the shift towards more student-driven, interactive learning methods, but our randomized controlled trial with a within-subject design provides a more robust comparison of these two methods within the same cohort, offering new insights into their relative effectiveness, demonstrating comparable benefits at initial exposure and suggesting potential cumulative advantages of role-playing over repeated sessions, rather than definitive superiority of one method. Furthermore, Hai Hu et al. [17] demonstrated that a role-playing computer game designed to strengthen clinical-decision making and situational awareness led to improved test performance and increased motivation among pre-clinical medical students, highlighting the educational value of immersive, experiential learning strategies. In parallel, our findings indicate that traditional, non-digital role-playing can also support and enhance student learning within a medical biochemistry PBL context. While Hai Hu et al. employed technology-driven disaster scenarios to create immersive learning environments, our study emphasizes a human-driven, face-to-face role-playing approach focused on conceptual understanding and application of biochemistry principles. Student feedback in our study suggested that role-playing was engaging and facilitated comprehension of complex topics, accompanied by significant improvements in post-test scores. Together, these findings suggest that both digital and traditional role-playing approaches are effective in promoting engagement and learning, with traditional role-playing offering a practical and pedagogically valuable alternative in foundational subject such as medical biochemistry, where direct interaction and guided discussion may be particularly beneficial. Sakamoto et al. [18] reported that students exposed to team-based learning [TBL] demonstrated significantly higher knowledge retention than those receiving lectures immediately after the intervention, although this advantage was not sustained at 30-day follow-up. In a comparable manner, our randomized controlled trial demonstrated significant short-term improvements in post-test performance following both case-based learning and role-playing sessions, indicating the effectiveness of student centered instructional approaches in medical biochemistry education. While unadjusted analysis in our study showed greater post-test scores and learning gains during several role-playing sessions, adjusted analysis restricted to the pre-crossover phase did not demonstrate a statistically significant between-arm difference at initial exposure. Together, these findings suggest that role-playing may support short-term engagement and learning gains over repeated exposure rather than confer immediate superiority, aligning with evidence from TBL-based interventions that emphasize active participation and collaborative learning. Both approaches were well received by students and reinforce the value of interactive pedagogies in promoting early learning outcomes.

In summary, our study is the first to directly compare role-playing and case studies in the context of medical biochemistry education using a randomized controlled trial (RCT) design, filling a critical gap in the existing literature. While previous studies have explored the effectiveness of case-based learning and role-playing in various contexts, our research uniquely examines both methods within the same cohort of medical students. We address the gap in understanding the comparative impact of these interactive teaching methods in medical biochemistry education. Our findings provide novel insights, demonstrating that both case studies and role-playing are effective problem-based learning strategies for improving learning outcomes in medical biochemistry. While unadjusted analysis suggested greater learning gains with role-playing over repeated exposure, the primary adjusted analysis showed comparable effectiveness of both methods at initial implementation. These findings indicate that role-playing may offer incremental benefits over time rather than clear early superiority. Overall, the results support the integration of active learning approaches to enhance student engagement and understanding in medical education, an area that remains underexplored.

### Study Limitations

While this study provides valuable insights into the comparative effectiveness of case studies and role-playing in medical biochemistry education, several limitations need to be considered. First, the study was conducted within a single institution, limiting the generalizability of the findings to other educational contexts or populations. Second, although the final analysed sample exceeded the pre-specified sample size requirement, it may not fully represent the diverse learning styles and preferences of all medical students. Although 150 students were initially enrolled and randomized, only 45 students completed all intervention sessions and assessments and were included in the final analysis. The attrition was primarily due to absenteeism during scheduled teaching sessions and assessments and may have introduced selection bias and may limit the generalizability of the findings. Nevertheless, the final analysed sample exceeded the minimum sample size calculated a priori (n=39). Furthermore, because the analysis was restricted to participants who completed all intervention sessions and assessments, the findings should be interpreted in the context of a complete-case analysis. In addition, the final analysed sample comprised predominantly female participants (84%). Although the within-subject randomised crossover design ensured that each participant received both instructional methods, thereby minimizing the potential influence of gender on the comparison between interventions, the predominance of female participants may limit the generalizability of the findings to student populations with a more balanced gender distribution. Finally, while the study focused on short-term outcomes, the long-term effects of both teaching methods on student retention and clinical practice remain unclear. Future studies could explore the impact of case studies and role-playing over an extended period to evaluate sustained knowledge retention and its application in real-world clinical settings.

### Future directions

Future research should aim to explore the long-term impact of role-playing and case study methods, particularly how these approaches influence clinical decision-making and problem-solving over time. Investigating the effects of these methods in diverse cohorts, such as students from different medical schools or those specializing in different fields, would help determine the generalizability of the results. Additionally, studies could examine how combining role-playing and case studies, or integrating digital technologies into these methods, can create a more hybridized, effective teaching approach. Exploring how these interactive methods can be adapted for online learning or in resource-limited settings would also contribute to understanding their scalability and accessibility in medical education.

## Conclusion

This study demonstrates that both case studies and role-playing, implemented as problem-based learning strategies, were effective in improving students’ understanding and application of medical biochemistry concepts. Significant pre- to post-test gains were observed with both methods, indicating their educational value. While unadjusted and pooled analyses, along with student feedback, suggested comparatively higher engagement and learning gains during several role-play sessions, the primary adjusted analysis restricted to the pre-crossover phase did not demonstrate a statistically significant difference between instructional methods at initial exposure. These findings suggest that both approaches are similarly effective when first introduced. Role-playing was perceived by students as particularly supportive of interactive learning, real-time decision-making, communication, teamwork, and confidence building, aligning well with AETCOM competencies. Case studies were consistently valued for their relevance, structure, and support for analytical reasoning and application of theoretical knowledge. Overall, the results indicate that role-playing may offer incremental and cumulative learning benefits with repeated exposure rather than clear superiority at the outset. Combining case studies with role-playing may therefore provide a complementary and balanced PBL framework. Future studies employing longitudinal designs and advanced analytical models are warranted to better elucidate time-dependent and cumulative instructional effects in medical education.

## Data Availability

All relevant data are within the manuscript and its Supporting Information files.

## Acknowledgement

The authors would like to sincerely thank the Phase I MBBS students who voluntarily participated in this study and actively engaged in the PBL sessions. We acknowledge the support of Department of Biochemistry faculty and supporting staff for their assistance in conducting PBL session. We are grateful to the institutional authorities-Dean, Medical Director and the Management Trustees of Sukh Sagar Medical College and Hospital, for granting permission to conduct this educational research.

